# The acceptability of being trained to deliver online, group-based Acceptance and Commitment Therapy to stroke survivors: the experience of third-sector practitioners

**DOI:** 10.1101/2024.10.08.24315102

**Authors:** Hannah Foote, Audrey Bowen, Sarah Cotterill, Emma Patchwood

## Abstract

**Purpose:** The Wellbeing After Stroke study (WAterS) co-developed a nine-week, online, group-based intervention for stroke survivors, informed by Acceptance and Commitment Therapy (ACT), to be delivered by third sector practitioners. This study explored practitioners’ perceptions of the acceptability of the training and their views on delivering the intervention.

**Materials and methods:** Semi-structured interviews completed with practitioners after training, but before intervention delivery had begun. Interview schedule guided by the Theoretical Framework of Acceptability. Template Analysis used to inductively and deductively interpret the data.

**Results:** All eight WAterS-trained practitioners were interviewed. Five main themes were generated. Practitioners were motivated to deliver a stroke-specific therapy. Practitioners reported that training was understandable and that experiencing ACT during training benefitted practitioners’ own wellbeing and increased their preparedness for intervention delivery. Previous experience affected their confidence to deliver. Practitioners expected the therapy to be acceptable to many stroke survivors. The online group context was expected to be beneficial, although they foresaw challenges in remotely facilitating groups with diverse accessibility needs.

**Conclusion:** It is acceptable to upskill a third-sector workforce to deliver a protocolised ACT-informed intervention to stroke survivors, potentially enabling greater reach of much needed psychological support.

## Introduction

Mental health issues are common after stroke ^1–3^, and support for these is the number one research priority for life after stroke ^4^. Access to clinical psychologists is limited ^5^, and stepped/matched care models for psychological interventions have been suggested as a means of involving the wider workforce ^6,7^. Acceptance and Commitment Therapy (ACT) ^8^ is a trans-diagnostic, third-wave, cognitive behavioural therapy with an expanding evidence base in supporting psychological difficulties post-stroke ^9–15^.

The Wellbeing After Stroke (WAterS) feasibility study ^16^ co-developed and demonstrated the feasibility of a protocolised, nine-week, group-based ACT-informed intervention, and accompanying training course, delivered online using Zoom. A third-sector workforce of practitioners employed by a UK national charity, the Stroke Association, without clinical training or previous experience of ACT, were trained and supported to deliver the intervention. The training course consisted of four half-day group sessions delivered by a clinical neuropsychologist (Geoff Hill, WAterS Clinical Lead) and the WAterS Principal Investigator (Emma Patchwood), and was accompanied by a practitioner handbook which included scripts to guide delivery. Each intervention group was run by two practitioners, a lead and a support. Practitioners received weekly supervision from a clinical neuropsychologist alongside delivery. A detailed description of the intervention and training is available^16^.

Examining the acceptability of an intervention at the feasibility stage of a study can inform modifications to improve the design of future research and implementation ^17,18^. The Theoretical Framework of Acceptability (TFA)^19,20^ defines acceptability as “the extent to which people delivering or receiving a healthcare intervention consider it to be appropriate, based on anticipated or experienced cognitive and emotional responses to the intervention”^20^, and posits seven components of acceptability, including burden, intervention coherence and perceived effectiveness. See Supplemental material S3 for the seven TFA components and their definitions.

This paper explores practitioners’ views on the acceptability of the WAterS practitioner training course and practitioners’ anticipated acceptability of delivering the WAterS intervention to stroke survivors.

## Materials and Methods

Ethics approval was secured from the University of Manchester Research Ethics Committee (ref 2021-11134-18220). This study is reported using the COnsolidated criteria for REporting Qualitative research (COREQ) checklist ^21^ (see Supplemental material S1).

This qualitative study used semi-structured, one-to-one interviews and took a ‘limited realist’ position ^22^, to recognise the subjectivity of the practitioners and researchers, while drawing on theory and assuming that findings have the potential to have wider relevance.

All the practitioners recruited and trained for the WAterS study were invited to take part in this study. Recruitment and consent were carried out online by members of the WAterS research team prior to the start of the practitioner training course.

The eligibility criteria were:

- Employed as frontline practitioners by the Stroke Association for at least 6 months
- Capacity to participate with clearance and support from line manager
- Experience and knowledge of facilitating groups of stroke survivors
- Willing to be trained and adhere to research procedures

### Study materials

The development of the interview schedule was informed by the Theoretical Framework of Acceptability and addressed each of the seven theorised components of acceptability ^20^, in relation to questions about both the practitioner training course and the anticipated delivery of the WAterS intervention groups (see Supplemental material S2 for the full interview schedule). Questions were piloted for clarity with a third party, a colleague with a similar professional background to the practitioners.

### Data collection and processing

All interviews were conducted by the first author (PhD student), who had previous experience of qualitative interviewing and was known to the practitioners as a member of the WAterS research team. The interviews took place after the practitioner training and before the start of the WAterS intervention delivery, via Zoom, in a private location with no-one else present. Interviews were recorded (audio and video) and securely stored. Field notes were taken as necessary. The recordings were transcribed verbatim and checked for accuracy.

### Data analysis

Data were managed using NVivo software. Findings were thematically analysed using Template Analysis ^23^, which allows for both inductive and deductive analysis and use of *a priori* themes. The analytic process included eight procedural steps ^23,24^:

1. Identified *a priori* themes: the seven TFA components
2. Read through all transcripts to familiarise with the data
3. Coded the data by *a priori* and themes inductively generated by researchers
4. Produced initial template of themes, grouped as either top-level or sub-themes
5. Applied initial template to full data set and re-defined, modified and collapsed top-level and sub-themes
6. Quality checks were carried out, and the template further revised:

a. Two authors separately coded one transcript according to the template, then compared and discussed codes to facilitate reflections on the analysis
b. Feedback was given on initial data analysis by a WAterS-specific Patient, Carer and Public Involvement (PCPI) advisory group
7. Template of themes finalised
8. Final template applied to full data set.

## Results

All eight participants who attended the practitioner training course were interviewed in July 2021, one-to-two weeks following the end of the training course and two-to-three weeks prior to delivering the WAterS intervention. All were female, with a mean age of 53 (SD: 7.63). The mean number of years working for the Stroke Association was 6 (range 1-15). Five participants were trained as lead practitioners and three as support practitioners. Due to Stroke Association workforce reorganisation, the recruited lead practitioners all had a UK Level Four counselling qualification although this was not a requirement of the role. The mean interview length was 54 minutes, ranging from 45–65 minutes.

### Findings

The final template used for analysis consisted of five study specific themes developed by researchers - see Supplemental material S3 for the iterative development of templates used during analysis. These five themes each contain data relevant to both of the two research objectives (the acceptability of the received training and the anticipated acceptability of delivering the intervention), as practitioners’ views on these were closely linked. Each of the five study specific themes relate to multiple TFA components, with all seven TFA themes being present in the data. Table 1 gives an overview of findings, and shows how these findings relate to the TFA components.

**Table 1:** An overview of key findings by theme, with reference to the TFA components.

| Theme | Key points (*"Relevant TFA component/s"*) |
| --- | --- |
| Motivation: emotional support is important and lacking for stroke survivors | Practitioners believed in the importance of mental health support for stroke survivors (*"Ethicality"*)<br>Practitioners felt positive and motivated to deliver the intervention as they perceived it as needed (*"Affective attitude"*)<br>Practitioners felt that taking part in the training and intervention was time-consuming and effortful, but this burden was reduced by motivation (*"Burden", "Opportunity costs"*) |
| Experiencing ACT is impactful | The practitioners personally benefitted from ACT during the training and therefore perceived the intervention as potentially effective for stroke survivors (*"Perceived effectiveness [of training and intervention]"*)<br>Experiencing ACT during training increased the practitioners' understanding of how the intervention works (*"Intervention [and training] coherence"*) |
| Best-fit: accessibility and structure of the training and intervention | The handbook was well-organised enabling the practitioners to focus on course content (*"Burden"*)<br>The structure of the training (four half-days with breaks) was acceptable and enabled time for reflection (*"Burden", "Perceived effectiveness [of training]"*). Some would have liked longer breaks (*"Burden"*)<br>Some training sessions overran and there was weekly home practice which increased the time commitment (*"Burden", "Opportunity costs"*)<br>The training used various methods and role-play which was perceived as supporting learning and engagement (*"Perceived effectiveness [of training]"*)<br>Practitioners felt the provided scripts would support them to deliver the intervention as intended, but were concerned it would make their delivery less responsive (*"Perceived effectiveness [of the intervention]"*) |
|  | • Practitioners considered the barriers the stroke survivors may face in engaging with the intervention and were concerned about how the intervention being time-limited would impact the stroke survivors (*“Perceived effectiveness [of the intervention]”*)<br>• Practitioners felt the client handbook (provided to the stroke survivors) would support accessibility (*“Intervention coherence”*)<br>• Practitioners thought the online modality would increase accessibility for some stroke survivors, but may add challenge to delivery (*“Burden”*) |
| Influence of previous experience and the need to clarify expectations | • Previous experience impacted on the practitioners’ understanding of the training course content (*“Intervention [training] coherence”*) and their confidence in their ability to deliver the intervention (*“Self-efficacy”*)<br>• Practitioners were varied in how confident they were about delivering a new intervention. The weekly supervision sessions were perceived as supporting learning (*“Self-efficacy”*)<br>• Practitioners felt they would need to spend time and effort independently preparing for delivery which they had not all anticipated (*“Burden” and “Opportunity costs”*)<br>• Clarifying expectations would improve the training course (*“Perceived effectiveness [of training]”*) and increase confidence (*“Self-efficacy”*) |
| Group relationships are important and challenging | • Receiving training in a group was reported to support learning (*“Perceived effectiveness [of training]”*)<br>• Practitioners enjoyed the relaxed atmosphere during training (*“Affective attitude”*)<br>• Practitioners anticipated that delivering the intervention in groups could be beneficial to stroke survivors, but that there was potential for stroke survivors to compare themselves negatively to others (*“Perceived effectiveness [of the intervention]”*)<br>• Facilitating the groups’ contributions was seen as an effortful aspect of delivery (*“Burden”*) |

Findings with exemplifying quotes will now be presented according to the five study specific themes.

#### Motivation: emotional support is important and lacking for stroke survivors

Practitioners came to the WAterS training with an existing belief in the importance of supporting the mental health of stroke survivors. They all recognised the need for more emotional support for stroke survivors, based on their experiences: “A huge proportion of people that we work with really struggle with their mental health […] and there’s a huge […] gap out there […]. It’s difficult for people to get support” [ID08]. Therefore, while taking part in the WAterS training and intervention delivery was seen as a big commitment in terms of time and effort, the practitioners were motivated to do so in order to increase their ability to best meet stroke survivors’ needs: “For many clients, they have so many diverse challenges, the […] more we learn, the more we can offer and the […] better outcomes for them in the long run” [ID02].

Practitioners were motivated to support stroke survivors with more severe cognitive and communication difficulties as they felt a particular lack of services for this group. Therefore, there was some disappointment that WAterS would not be fully accessible for all stroke survivors. However, practitioners valued the adaptations that had been made to enhance accessibility: “If somebody had severe aphasia, it would be not […] fully accessible, but […] I was quite impressed with the fact that language and cognition had been considered […] and that the exercises would be accessible to the majority” [ID08].

As practitioners were so motivated to provide appropriate support to stroke survivors, the slow pace of research was a cause of some frustration, with one practitioner saying: “I would just love to be able to actually just go out and deliver this without it being a pilot study” [ID05].

#### Experiencing ACT is impactful

Practitioners perceived that they had gained personal benefit from carrying out the ACT-informed exercises as part of their training: “I’ve been using it more in my life; noticing things, changing how you think about things, being present in the moment” [ID01]. This led them to feel hopeful about the effectiveness of the WAterS intervention for stroke survivors: “If things work for you […] then you can see how it’s going to work for other people” [ID02].

The practitioner training was delivered in an ACT-consistent manner, with one practitioner stating that the trainer (WAterS Clinical Lead) “sort of lived it, the ACT” [ID03]. Experiencing ACT supported the practitioners to understand the intervention: “The experiential exercises we did are really helpful and really enable us to empathise and to mirror what the stroke survivors is going to be experiencing” [ID05]. Some practitioners demonstrated their belief that the approach may be effective by reporting that they’d started integrating ACT into their usual work and informally when supporting family and friends: “Even in my one-to-one practice with my clients, I can […] feel it creeping in” [ID04].

The practitioners all demonstrated an understanding of how ACT works, referring to key components of the ACT model and applying this to post-stroke experiences. Two components, mindfulness and values identification were seen as particularly important and relevant to life after stroke: “It’s […] inviting people to reassess their values post-stroke because maybe their views will have changed considerably” [ID05].

Practitioners perceived the inclusion of goal-setting activities as important in enabling the stroke survivors to identify practical ways to move forwards, and the weekly home practice as important in supporting the stroke survivors to embed ACT in their lives: “The homework that they’ve got to do […] I think that’s really good because quite often in therapy people go from week to week and don’t do the work in between” [ID01].

Practitioners understood that WAterS was designed to engage people through the use of various strategies and that not everything in the intervention would be applicable or relatable to all. However, one practitioner stated that this ethos hadn’t been clear at first and felt this would be important to clarify to participating stroke survivors: “I’ve realised […] this is not a compliance exercise […], this is presenting them with a lot of options and some of them will click […] and it’s okay to say, this one’s not working for me, I’m going to sit this one out” [ID06].

#### Best-fit: accessibility and structure of the training and intervention

Practitioners described their training as easy to follow and the practitioner handbook as well-organised, enabling them to focus on the course content: “The handbook was neatly done and […] in one folder. That was just so key for me because I would be flapping about with that” [ID03]. The practitioners were broadly satisfied with the structure of the training course: “It was nice in four sessions to break it down […] it was nice that we could reflect each week” [ID02]. Each training session included necessary breaks, but some felt these breaks were too short. The time commitment was a burden for some participants, particularly if training sessions overran and when trying to fit in the weekly practice at home.

The practitioner training included several different methods to support learning, which was reported to improve engagement and maintenance of attention. However, one practitioner felt that there were too many short exercises and would have preferred focusing for longer on fewer exercises. Opportunities to role-play delivery were reported as helpful in increasing the practitioners’ confidence: “I was a bit apprehensive about delivering some of the guided breathing […] but we practised it in the [training] group and I feel a bit better” [ID07].

In terms of delivering the WAterS intervention, some practitioners expressed concern that the nine-week structure of the WAterS intervention may not provide enough time to adequately address issues arising during the course: “You don’t want to open something up and then leave people dangling at the end of it” [ID03]. Practitioners felt that the scripts provided to guide intervention delivery would be useful: “Because if it wasn’t [scripted], I’d be ad libbing and […] maybe straying away.” [ID03]. However, practitioners wanted to ensure they were responsive to the stroke survivors in the group: “I don’t want to talk at them, I want to […] be with them” [ID01]. Practitioners noted that the non-scripted aspects of the course may be the most challenging to deliver: “The big challenge is about trying to respond to people in an ACT-coherent manner when you’re on the spot” [ID08].

In preparing for delivery, practitioners considered the possible barriers stroke survivors may have in engaging with the intervention and how they could facilitate their engagement: “I know IT [information technology] is a big one [barrier] and […] confidence and speech and fatigue and concentration […] then as a facilitator, how am I going to manage that?” [ID01]. Practitioners felt that the accessibility of the WAterS intervention and the client handbook (provided to stroke survivors at the start the intervention) was increased through the use of visuals and simple language, with one practitioner suggesting ways the accessibility of the client handbook could be increased: “Making it [the client handbook] more distinctive in terms of sections or colour […] to […] make things slightly more visible and scannable” [ID06].

The positives and negatives of the online modality were considered by the practitioners. They felt it could increase accessibility for people with physical limitations or living in remote areas, but that the inclusion of technology may be an added barrier for some: “When I started to use Zoom, you couldn’t really focus on […] what you were trying to say, you were so worried […] that it was going to go wrong […], I can imagine it would be amplified in somebody who’s had a stroke” [ID03].

#### Influence of previous experience and the need to clarify expectations

The lead practitioners had previous counselling training and stated that this helped them to understand the WAterS training content and to integrate the information with their existing knowledge. The support practitioners needed more time to develop their understanding: “I have no background in counselling at all […], it was completely new to me […]. At the beginning [it] very much seemed like a big knot of things and then gradually it all started to come out into some sensible series of conversations” [ID06]. In relation to delivery, lead practitioners reported that their counselling skills would be very important, with some feeling they would be essential: “If I’m delivering that […] I really will very much have my counsellor’s hat on and be prepared for the unexpected and […] to hold and maintain the people that I’m working with.” [ID04]. Support practitioners felt that they required more support in dealing with stroke survivors’ difficult emotions that may arise during the intervention.

The practitioners had varying levels of confidence about delivering the intervention. As well as previous experience, practitioners referred to their usual approaches to doing new things. Some practitioners reported feeling comfortable to learn alongside delivery, supported by the weekly clinical supervision sessions: “it is good that we’ve got that supervision with [WAterS Clinical Lead] […], so if there’s anything that’s missing, I think we’ll learn as we go along” [ID02]. Whereas other practitioners expressed anxiety or worry about the prospect of delivering the sessions: “My fantasy is a case of getting stuck on all the words and […] deviating from the script and not being able to get back to it. All these fears that go through your head. And I’m sure some coordinators [practitioners] may not have those anxieties” [ID05].

While all practitioners stated that they would do independent preparation prior to delivering the course, this preparation time was particularly important to those less confident and/or with less experience. Practitioners would have liked this time to have been protected from the outset: “I will spend quite a lot of time preparing and it’s obviously not something […] with our managers that was discussed that we would need to do and maybe a bit underestimated” [ID07].

Practitioners expressed the importance of clarifying expectations as part of the training course. The support practitioners communicated some uncertainty about their remit and would have liked the training to focus on the purpose of their role. The lead practitioners would have liked more clarity on the processes that they would hold responsibility for, including risk management procedures. Some lead practitioners would have liked more training on the practicalities of delivering the intervention: “I would have liked […] more time spent […] on delivering each session […], maybe the challenges that you might have in that session […]. Even if the [training] sessions had been half an hour longer, it would have been really helpful” [ID05].

The trainer (WAterS Clinical Lead) mirrored delivery of the intervention to the practitioners during the training. While this was seen as useful for learning, one practitioner stated that the trainer had set a very high bar, making it more difficult to identify what the expected quality of delivery might be: “I thought the delivery of them [the WAterS training sessions] was […] brilliant, almost so good that it made me anxious about delivering it myself” [ID07].

#### Group relationships are important and challenging

The practitioners valued the group aspect of the practitioner training course, they reported learning from each other and relating to each other’s experiences within the group. The size of the training group (eight practitioners and two trainers) ensured everyone had a chance to contribute. The practitioners reported enjoying the informal, relaxed atmosphere created by the trainers and felt comfortable asking questions. The practitioners appreciated that the trainers recognised and trusted the practitioners’ own expertise: “We’re doing this all the time, talking to our clients who’ve had strokes and their families, and it was nice that you guys listened to what we were saying as well” [ID01].

In terms of delivery, the practitioners anticipated the importance of the relationships that they would form with the stroke survivors: “Feeling that somebody understands, somebody’s trying to help, somebody’s walking through it with them, I think they can benefit” [ID02]. The practitioners reflected on group aspect of the WAterS intervention and viewed peer support as a potential benefit: “I’ve seen amazing things happen with peer support […] motivating each other and unpredictable effects from that” [ID06]. However, one practitioner reflected on the possibility that stroke survivors may compare themselves negatively to others in the group: “They might feel it was worse than the others and start comparing themselves with their peers on that Zoom.” [ID02].

Practitioners anticipated that facilitating and containing the groups’ contributions would be a challenging aspect of delivery. They felt that intervention group sizes would need to be fairly small in order to ensure there was time for all stroke survivors to feedback: “I think the challenge comes from managing the feedback from participants [stroke survivors] and being able to juggle that with getting through the amount you have to” [ID05]. One practitioner considered whether there was merit in active grouping of people, to tailor delivery to the needs of the group: “In the longer term, it could well be that it’s more suited to put people of a similar ability together, so maybe you can go at a slower pace.” [ID05].

The practitioners all had experience of running groups but felt that the online nature of the groups would increase effort in delivery and that communication may be more challenging via this medium. This was a particular concern in conjunction with delivering potentially emotionally confronting content, and the possibility of emotionalism ^25^ experienced by stroke survivors: “it’s so easy if you’re in a room with somebody to pick up on that body language […] the eyes, the twiddling the fingers […]. It is having to use slightly different skills to try and pick up what you can on the screen” [ID02]. Practitioners felt that they had benefitted from observing the trainer (WAterS Clinical Lead) facilitating their discussions during the training course, but that there would be different challenges in facilitating groups of stroke survivors, with one practitioner stating that it would be helpful to observe the trainer delivering the intervention to stroke survivors.

## Discussion

The WAterS training course was broadly acceptable and understandable to the practitioners. The training provided the practitioners with an experience of Acceptance and Commitment Therapy, which led to perceived benefits in their own wellbeing, as well as preparing them to deliver the online intervention to groups of stroke survivors. Previous experience impacted on both understanding of the training and confidence to facilitate the WAterS intervention. Clarifying expectations during training and the provision of additional preparation time would increase preparedness for delivery. Prior to delivery, practitioners reported feeling highly motivated to facilitate the intervention and perceived the intervention as being accessible and relevant for many stroke survivors. The online group-based delivery mode was perceived as adding challenge, but potentially beneficial in increasing access and support for stroke survivors. The Theoretical Framework of Acceptability was a useful tool in ensuring a comprehensive investigation of acceptability.

To our knowledge, this is the first paper on ACT for stroke which considers the perspective of the practitioner in-depth. It is promising that the practitioner training course was perceived to be understandable to a third-sector workforce without previous ACT expertise, as previous studies of protocolised ACT interventions for stroke survivors have been led by qualified psychologists ^9,26,27^ or psychological therapists ^12^. The practitioners perceived benefits to their own wellbeing from the experience of ACT, in line with previous research indicating that attending ACT training can have personal benefits ^28^. A previous qualitative study exploring stroke survivor views of an ACT-informed intervention^29^ found that stroke survivors valued facilitators being knowledgeable and authentic, and the results of the present study indicate that the WAterS training provided opportunities to develop these skills.

There are strengths and limitations in the use of the Theoretical Framework of Acceptability (TFA) in this study ^20^. The use of the TFA during data collection supported us to be comprehensive in investigating acceptability and all components of the TFA were relevant to the data collected in this study (see Table 1). Validation of the TFA is ongoing and therefore we also used inductive methods of analysis. When applying the TFA to the data, overlaps in the components were noted, for example, if something was identified as a “Burden” it often also incurred an “Opportunity cost” (see Table 1). Therefore we found that developing study specific themes best reflected the data. Prospectively exploring the acceptability of delivering the intervention enabled identified issues to be addressed prior to delivery.

A strength of the study is that all practitioners involved in WAterS were interviewed, so no sampling was required. However, there are some key factors that may limit the generalisability of findings to a wider potential workforce. All practitioners had volunteered to take part in the WAterS study, and the lead practitioners in this study were qualified counsellors, which we had not anticipated as it was not an eligibility criteria. Furthermore, all practitioners were white females, and whilst this is largely representative of the demographics of the Stroke Association’s workforce, it may limit their ability to best-serve the UK’s ethnically diverse population of stroke survivors ^30^.

Interviews were carried out within two weeks of the practitioner training, to support recall of the experience, and one-to-one interviews allowed the in-depth exploration of practitioners’ views. The practitioners were encouraged to be open and honest in their interactions with the interviewer, however the interviewer was a member of the research team and this may have impacted on the practitioners’ responses.

Throughout the study, authors engaged in reflexivity to increase awareness of subjectivity and think critically about analysis. In line with the limited realist position, quality checks were carried out during data analysis, including independent coding to prompt discussion and reflection on analysis. The WAterS PCPI group were consulted regarding preliminary results and analysis, and they identified areas that required further clarification and ensured the language used was clear and respectful.

The results of the present study indicate that the WAterS training and delivering the intervention were acceptable to practitioners, with other WAterS outputs highlighting that the practitioners were able to deliver the intervention with high fidelity to the protocol and that the intervention was broadly acceptable to the stroke survivors who received it ^31,32^. However, online group support delivered by this workforce may not be appropriate or accessible to all stroke survivors. Future research should investigate suitability of the growing body of ACT-informed interventions for stroke survivors ^9–15^ to different people within the stroke population. To reduce health inequalities, future research should be co-developed with under-served populations, including minoritised ethnic groups and stroke survivors with more severe cognitive and language difficulties. The Wellbeing After Stroke-2 (WAterS-2) study^33^ began in October 2023 to address these areas.

In conclusion, this study showed that it is acceptable to upskill a third-sector workforce to deliver a protocolised ACT-informed online intervention to stroke survivors and this could enable greater reach of much needed psychological support to stroke survivors. This study provides useful insights which may support the development of future psychological therapies designed for delivery by a third-sector workforce, such as the importance of providing experiential training, valuing the contributions of the workforce, clarifying roles, ensuring procedures are clearly specified, and protecting sufficient time for preparation.

## Supporting information

S1 COREQ checklist

S2 interview schedule

S3 iterative development of themes

## Acknowledgments

This independent research was funded by the University of Manchester Research Impact Scholarship and a Stroke Association Postdoctoral Fellowship (Ref SA PDF 18100024). The funders had no role in study design, execution, analysis or results interpretation and the views expressed are those of the author(s) and not necessarily those of the funders.

The authors extend thanks to the participants, the members of the WAterS Patient, Carer and Public Involvement group and the Clinical Lead for the Wellbeing After Stroke study (Dr Geoff Hill, Clinical Neuropsychologist).

## Data Availability Statement

The data that support the findings of this study may be available, on reasonable request from the corresponding author, for five years following publication.

