## Supplementary material for "The acceptability of being trained to deliver online, group-based Acceptance and Commitment Therapy to stroke survivors: the experience of third-sector practitioners": S2 interview schedule

Supplemental material S2: semi-structured interview schedule

**Prospective acceptability of the WAterS intervention**

| |
|---|
| **Opener**: Can you tell me a bit about why you wanted to be part of this project? |
| **Affective attitude (how a person feels about the intervention)** |
| Now you have completed the training, how do you feel about the WAterS therapy?<br><br>What do you like about the therapy?<br><br>What don't you like about the therapy?<br><br>*Optional: How does this compare to other therapies you have delivered?*<br><br>How do you feel about delivering the therapy to people with cognitive or communication difficulties?<br><br>How do you feel about the remote/online delivery? |
| **Ethicality (good fit with value system)** |
| How well does the therapy fit with your personal values? |
| **Intervention coherence (understanding of the intervention and how it works)** |
| What is your understanding of the purpose/aims of therapy? What do you think it's trying to achieve?<br><br>How familiar does the protocol feel at this stage?<br><br>In order to achieve this, what do <u>you</u> think is important for you to do as the trainer?<br><br>What do <u>you</u> think are the key strategies/activities in the intervention? |
| **Perceived effectiveness (perception of whether intervention will achieve its purpose)** |
| How do you feel about whether the therapy will improve wellbeing for stroke survivors?<br><br>Which (if any) specific aspects of the therapy/its delivery do you think will contribute to achieving its aims?<br><br>What do you think might get in the way it being effective?<br><br>What challenges/barriers do you anticipate? |

| Burden (perceived effort in participating) |
|---|
| Thinking about what you know about the therapy so far, how do you feel about the effort that will be required from you to deliver it?<br><br>*Optional: How does this compare to other interventions you deliver?* |
| **Opportunity costs (what must be given up to engage in the intervention)** |
| Is there anything you are going to have to give up in order to deliver this therapy? How do you feel about that? |
| **Self-efficacy (confidence in performing the required behaviours)** |
| How confident do you feel about delivering the therapy?<br><br>Which parts of the intervention do you feel most/least confident in delivering? (probe for examples)<br><br>Can you tell me ways that we could support you to feel more confident? (caveat - limited in this study) |
| **Other** |
| Any further recommendations for us?<br><br>Anything you would change about the therapy? *(make it clear we won't necessarily be able to make changes)*<br><br>Anything else you would like to add? |

2. **Retrospective acceptability of the training**

| Affective attitude (how a person feels about the training) |
|---|
| How did you feel about the training overall?<br><br>What did you like/dislike about the training? |
| **Ethicality (good fit with value system)** |
| How well did the training course fit with your personal values? |
| **Perceived effectiveness (perception of whether training has achieved its purpose)** |
| To what extent has the training prepared you to deliver the intervention? |

| |
|---|
| Can you tell me ways the training could be improved to better support you? |
| **Self-efficacy (confidence in performing the required behaviours)** |
| How could we improve the training to help you feel more confident in delivery of the therapy?<br><br>In terms of the training course itself, how confident did you feel taking part in this?<br><br>Were there any particular parts of the training that you felt least/most confident in participating in? |
| **Intervention coherence (understanding of the training and how it works)** |
| How easy or difficult did you find the training to understand?<br><br>Which parts were easy/difficult? |
| **Burden (perceived effort in participating)** |
| How do you feel about the level of effort that was required for you to engage in the training?<br><br>*Optional: How does this compare to other training courses you have been on?* |
| **Opportunity costs (what must be given up to engage in the training)** |
| Was there anything you had to give up to attend the training?<br><br>How do you feel about that? |
| **Other** |
| Any recommendations for us?<br><br>Anything you would change about the training? *(make it clear we won't necessarily be able to make changes)*<br><br>Anything else you would like to add? Please feel free to contact me by email if you think of anything else. |
