## Supplementary material for "The acceptability of being trained to deliver online, group-based Acceptance and Commitment Therapy to stroke survivors: the experience of third-sector practitioners": S3 iterative development of themes

### Iterative development of templates used for data analysis

#### Initial template one (*a priori* themes): The Theoretical Framework of Acceptability[1] components

| **Main themes (TFA components)** | ***TFA definitions***[1] |
|---|---|
| Affective Attitude | *How an individual feels about the training/intervention* |
| Burden | *The perceived amount of effort that is required to participate in the training/intervention* |
| Ethicality | *The extent to which the training/intervention is a good fit with the individual's value system* |
| Intervention coherence | *The extent to which the participant understands the training/intervention and how it works* |
| Opportunity costs | *The extent to which benefits, profits or values must be given up to engage in the training/intervention* |
| Perceived effectiveness | *The extent to which the training/intervention is perceived as likely to achieve its purpose* |
| Self-efficacy | *The participant's confidence that they can perform the behaviour(s) required to participate in the training/intervention* |

1. Sekhon M, Cartwright M, Francis JJ. Acceptability of healthcare interventions: An overview of reviews and development of a theoretical framework. BMC Health Serv Res [Internet]. 2017;17(1):1–13. Available from: http://dx.doi.org/10.1186/s12913-017-2031-8

**Template two – identifying sub-themes**

| **Main themes** | **Sub-themes** |
|---|---|
| Affective Attitude and Ethicality | Not just chalk and talk<br><br>Atmosphere<br><br>There is a need for support |
| Burden | Non-scripted facilitation is more effortful<br><br>Cognitive load<br><br>Managing an online group<br><br>Barriers for stroke survivors |
| Training & Intervention coherence | Accessibility<br><br>Impact of previous experience<br><br>Understanding how ACT works |
| Opportunity costs | Balancing commitments |
| Perceived effectiveness | Clarity of roles<br><br>Inclusion of ACT exercises supported learning<br><br>Implementing a scripted protocol<br><br>Relationships<br><br>Perceived suitability to stroke survivors |
| Self-efficacy | Pre-existing confidence levels<br><br>Less confident about the unscripted delivery |

**Template three – development of inductive main themes due to overlap in sub-themes across the TFA components**

| **Sub-themes** | **Inductive main themes** |
|---|---|
| There is a need for support<br><br>Barriers for stroke survivors<br><br>Accessibility | Motivation: emotional support is important and lacking for stroke survivors |
| Understanding how ACT works<br><br>Inclusion of ACT exercises supported learning<br><br>Not just chalk and talk | Experiencing ACT is impactful |
| Non-scripted facilitation is more effortful<br><br>Less confident about the unscripted delivery<br><br>Implementing a scripted protocol<br><br>Perceived suitability to stroke survivors<br><br>Barriers for stroke survivors<br><br>Accessibility<br><br>Balancing commitments<br><br>Managing an online group | Best-fit: accessibility and structure of the training and intervention |
| Impact of previous experience<br><br>Clarity of roles | Influence of previous experience and the need to clarify expectations |

| | |
|---|---|
| Pre-existing confidence levels<br><br>Balancing commitments<br><br>Cognitive load | |
| Atmosphere<br><br>Relationships<br><br>Perceived suitability to stroke survivors | Group relationships are important and challenging |

**Final template**

| **Main themes** |
|---|
| Motivation: emotional support is important and lacking for stroke survivors |
| Experiencing ACT is impactful |
| Best-fit: accessibility and structure of the training and intervention |
| Influence of previous experience and the need to clarify expectations |
| Group relationships are important and challenging |
